# COVID-19 and climate: global evidence from 117 countries

**DOI:** 10.1101/2020.06.04.20121863

**Authors:** Simiao Chen, Klaus Prettner, Michael Kuhn, Pascal Geldsetzer, Chen Wang, Till Bärnighausen, David E. Bloom

## Abstract

Visual inspection of world maps shows that coronavirus disease 2019 (COVID-19) is less prevalent in countries closer to the equator, where heat and humidity tend to be higher. Scientists disagree how to interpret this observation because the relationship between COVID-19 and climatic conditions may be confounded by many factors. We regress confirmed COVID-19 cases per million inhabitants in a country against the country’s distance from the equator, controlling key confounding factors: air travel, distance to Wuhan, testing intensity, cell phone usage, vehicle concentration, urbanization, and income. A one-degree increase in absolute latitude is associated with a 2.6% increase in cases per million inhabitants (p value < 0.001). The Northern hemisphere may see a decline in new COVID-19 cases during summer and a resurgence during winter.

**One Sentence Summary:** An increase in absolute latitude by one degree is associated with a 2.6% increase in COVID-19 cases per million inhabitants after controlling for several important factors.

## Main Text

Many inhabitants of the Northern Hemisphere hope that spread of severe acute respiratory syndrome coronavirus 2 (SARS-CoV-2) and therefore prevalence of coronavirus disease 2019 (COVID-19) will decrease when the weather gets warmer and more sunlight reaches the Earth’s surface in spring and summer. Many viral acute respiratory tract infections, such as influenza A and B, rhinovirus, respiratory syncytial virus, adenovirus, metapneumovirus, and coronavirus, are climate dependent and share the same seasonality *(1)*. Some viruses may have better stability in low-temperature, low-humidity, and low–UV radiation environments *(2, 3)*. Thus, an association between hot and humid climate conditions and slower spread of SARS-COV-2 is plausible.

However, as of yet little evidence supports this hypothesis *(4)*. On March 9, 2020, the World Health Organization (WHO) stated that “[f]rom the evidence so far, the COVID-19 virus can be transmitted in all areas, including areas with hot and humid weather” *(5)*. On April 7, 2020, the U.S. National Academies of Sciences, Engineering, and Medicine concluded that “[a]lthough experimental studies show a relationship between higher temperatures and humidity levels, and reduced survival of SARS-CoV-2 in the laboratory, there are many other factors besides environmental temperature, humidity, and survival of the virus outside of the host that influence and determine transmission rates among humans in the ‘real world’… with natural history studies, the conditions are relevant and reflect the real-world, but there is typically little control of environmental conditions and there are many confounding factors” *(4)*.

We use global data to examine the relationship between climatic conditions and the spread of COVID-19 controlling for several important confounding factors. We regress the prevalence of COVID-19 (logarithmically transformed) at the country level against the latitude of a country. Latitude captures every climate, because different latitudes on Earth receive different amounts of sunlight. The farther from the equator a country is located, the sharper is the angle of the sun’s rays that reach it, the less UV radiation it receives, and the lower the temperature is. Furthermore, latitude also affects humidity, because evaporation is temperature dependent *(6)*.

To control for key confounders, our analysis includes (i) data on air travel *(7)* and the distance of a country from Wuhan *(8)*—the original epicenter of the epidemic—to capture the transmission of SARS-CoV-2 to a country via different routes *(9)*; (ii) vehicle concentration *(10)* and urbanization*(7)* to capture differences in the transmission of SARS-CoV-2 within a country *(11)*; (iii) testing intensity *(12, 13)* to control for the vigor of a country’s COVID-19 response and for detection bias in cross-country comparisons *(14, 15)*; (iv) cell phone usage *(7)* to control for the speed at which information on behavior change for COVID-19 prevention travels within a country *(11, 16)*; and (v) income *(7)* to control for the availability of general and health systems resources to contain the spread of SARS-CoV-2 *(17–19)*. We imputed missing country covariate data using multiple imputation *(20)*.

**Fig. 1** and **Table 1** show our results. The farther a country is located from the equator, the more cases the country has in relation to the number of inhabitants. Consistent with our expectations, COVID-19 prevalence is higher in countries that are more open to air travel and have higher vehicle concentrations. In ordinary least squares (OLS) regression, in which we control for all seven potential confounding factors, an increase in the distance from the equator by one degree of latitude is associated with an increase of the prevalence of COVID-19 by about 2.6% (**Table 1, Model 5**). This result is highly significant and implies that a country that is located 1000 kilometers closer to the equator could expect 21 percent fewer cases per million inhabitants, other things equal (given that a degree of latitude translates on average into a distance of 111 kilometers). Since the change in Earth’s angle towards the sun between equinox and solstice is about 23.5 degrees, one could expect a reduction in cases per million inhabitants by 46 percent between two seasons. The coefficients of the added quadratic terms are insignificant at the 5% significance level, indicating that the relationship we are estimating is approximately linear (**Table S1**). As a robustness check, we further perform robust estimation based on the MM estimator, which is less sensitive to outliers. The size of the coefficient for latitude in the MM estimation is slightly higher than that in the OLS estimation and it remains highly significant (**Table S2**).

**Fig. 1.**
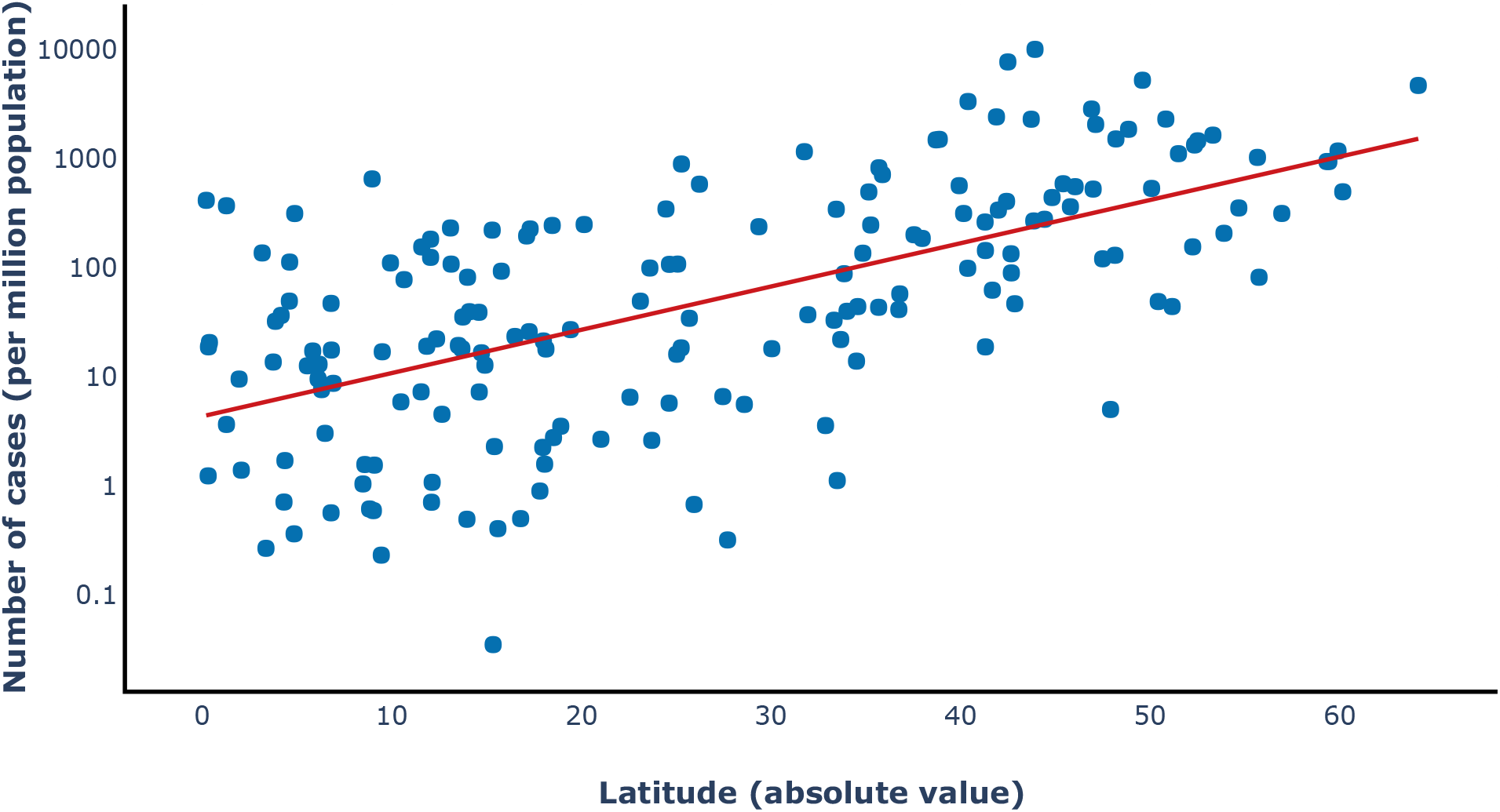
Scatterplot of the logarithm of cases per million inhabitants against absolute latitude in degrees for the full sample of countries.

**Table 1.**
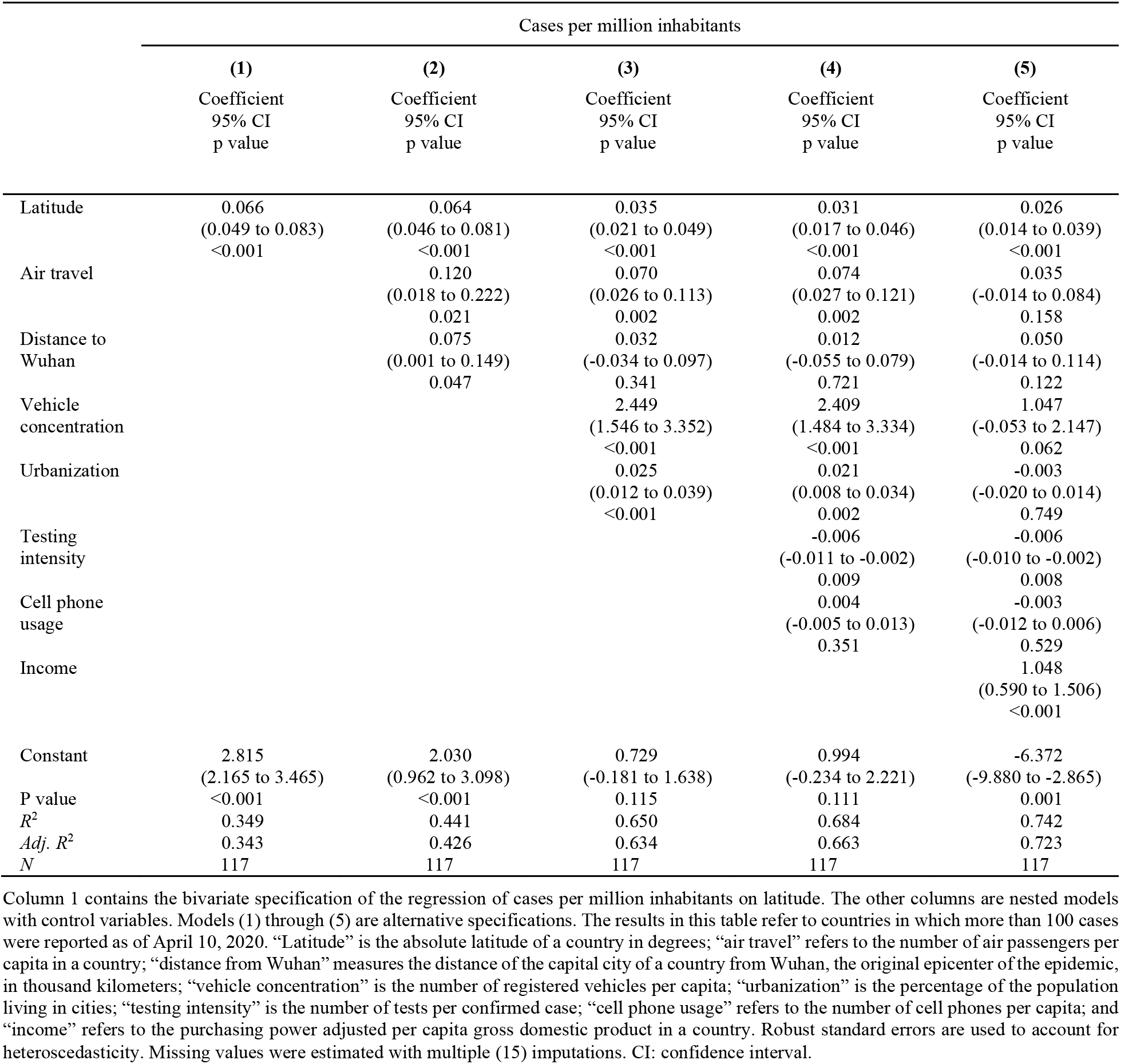
Results from Ordinary Least Squares regressions of COVID-19 cases per million inhabitants in a country on the country’s latitude and control variables.

Our results are consistent with the hypothesis that heat and sunlight reduce the spread of SARSCoV-2 and the prevalence of COVID-19. However, our results do not imply that the disease will vanish from the Northern Hemisphere during summer. Rather, the higher temperatures in summer are likely to support public health measures to contain SARS-CoV-2 to some extent. WHO’s warning that the virus spreads in all climates must still be taken seriously. Notably, the disease is likely to spread more easily in the Southern Hemisphere over the coming months as it enters fall and winter. Countries in the Southern hemisphere, such as Brazil, Indonesia, or especially South Africa, where the highest number of immunocompromised HIV-positive population live *(21)*, are less prepared to tackle the pandemic than the rich countries of the Northern Hemisphere.

Our analysis has several limitations. First, while our results are consistent with the hypothesis that higher temperatures and more intensive UV radiation reduce SARS-CoV-2 transmission, the precise mechanisms for such an effect remain unclear and may indeed constitute both biological and behavioral factors. For example, people might gather less in crowded indoor places if temperatures are higher, which would reduce transmission. Thus, future research at a later stage of the pandemic should aim at uncovering how the transmission of SARS-CoV-2 is impacted by changes in i) climatic factors such as heat and humidity, ii) geographic factors such as altitude and sunlight intensity, and iii) factors related to human behavior such as social interactions and pollution due to local economic activity at a more disaggregated level. Second, even though we include all countries worldwide for which data for this analysis were available, our final data set included only 117 out of the world’s 195 countries, mainly due to the fact that some countries have not yet surpassed the 100 case threshold. Third, while we strive to control for differential testing intensity using a recently compiled and frequently updated data set *(12, 13)*, the data on testing intensity could suffer from reporting biases and incomplete coverage of testing approaches. To the extent that testing intensity is a function of a country’s income, our analysis controlling for income (**Table 1, Model 5**) should reduce any biases these data limitations introduce. The fact that column (5) in **Table 1** contains a parameter estimate of latitude that is only slightly lower than the one in column (4) and still highly significant is reassuring in this regard. Finally, the distance to the equator only has the same effects to the south and the north at equinox. However, this would be a greater concern if we did the analysis closer to the solstices in summer or in winter. In addition, the effect sizes stayed rather stable over time. In earlier analyses of the data in mid-March, we found similar coefficient estimates.

In sum, we show that an increase in absolute latitude by one degree is associated with a 2–3 percent increase in cases per million inhabitants. Increasing temperatures and longer sunlight exposure during summer in the Northern hemisphere may to some extent boost the effectiveness of public health policies and actions to control the spread of SARS-CoV-2. Conversely, the threat of epidemic resurgence may increase during winter.

## Data Availability

All data are available in the main text or the supplementary materials.

## Acknowledgments

We would like to thank Ana Abeliansky, Ilan Gutherz, Catherine Prettner, Alfonso Sousa-Poza, and Alexander Zeuner for valuable comments and suggestions.

## Author contributions

SC, KP, TB, DB, and CW contributed to the study concept and design. SC and KP collected data, conducted data analysis, visualized results, and wrote the first draft of the manuscript. SC, KP, TB, DB, and CW contributed to literature review and the interpretation of the data. TB, DB, CW, PG, and MK critically revised the manuscript for important intellectual content. All authors approved the final version. The corresponding authors attest that all listed authors meet authorship criteria and that no others meeting the criteria have been omitted. **Competing interests:** Authors declare no competing interests.

## Data and materials availability

All data are available in the main text or the supplementary materials.

